## Supplementary material for "Impact of the COVID-19 pandemic on cognitive function in Japanese community-dwelling older adults in a class for preventing cognitive decline": S1 Table

**S1 Table. English translation of the relevant questions used in this study.**

(The questions are translated from Japanese)

**The questionnaire on lifestyle and thoughts during the class suspension.**

1. What type of anxiety did you experience during class suspension? (Check all that apply)

□ Worrying about physical condition

□ Getting sick (What are the symptoms?: )

□ Progression of forgetfulness

□ Weakening of lower body

□ Reduced food intake

□ Decreased conversation

□ Difficulty sleeping

□ Increased stress (What kind of stress is it?: )

□ (Other: )

2. What did you consciously try to do during class suspension? (Check all that apply)

□ Exercising (e.g., gymnastics, walks, etc.)

□ Working on cognitive training (e.g., puzzles, coloring book, etc.)

□ Having conversations

□ Eating a well-balanced diet

□ Living a well-regulated lifestyle

□ Engaging in hobbies (What kind of hobby?: )

□ Collecting information on dementia by television, radio, and newspaper

□ (Other: )

3. Questions about exercise

3.1. What kind of exercise did you do? (Check all that apply)

□ No exercise

□ Walk

□ Exercises in the classroom (spontaneously)

□ Exercises in the classroom (watching them on television)

□ (Other:___________________)

3.2. How many days a week did you exercise?

□ None

□ 1–2 days

□ 3–5 days

□ Almost every day

3.3. What was the average duration of one exercise session?

□ < 30 minutes

□ 30–60 minutes

□ > 60 minutes

4. Questions about cognitive training (e.g., puzzles, coloring book, etc.)

4.1. Did you perform any cognitive training?

□ No

□ Yes (What kind of cognitive training did you perform?: )

4.2. How many days a week did you perform cognitive training?

□ None

□ 1–2 days

□ 3–5 days

□ Almost every day

4.3. Did you work on the cognitive training worksheets provided by the town hall?

□ No

□ Yes

5. Questions about conversation

5.1. How often did you have a conversation?

□ Rarely

□ Once a week

□ Once every 2–3 days

□ Almost every day

5.2. How did you have the conversation? (Check all that apply)

□ Directly

□ Via telephone

□ Using an online communication tool

□ (Other: )

6. If the class is suspended again, what do you want to be aware of or hope for?

Please feel free to write down anything you want.

Note: The results of items that were similar to the other questions are not presented in the main text (questions whose results were not shown were 3.1, 3.2, 3.3, 4.1, and 4.2.). The results are shown in S2 Table. In addition, descriptive answers were also excluded from the analyses.
