## Supplementary material for "Impact of the COVID-19 pandemic on cognitive function in Japanese community-dwelling older adults in a class for preventing cognitive decline": S2 Table

**S2 Table. Comparison of questionnaire results between the cognitive decline and non-decline group (results not shown in the main text).**

|  | Decline group | Non-decline group | Unadjusted | | Adjusted | |
| --- | --- | --- | --- | --- | --- | --- |
|  | (n = 16) | (n = 72) | OR (95% CI) | P value | OR (95% CI) | P value |
| Type of exercise |  |  |  |  |  |  |
| No exercise | 1 (6.3) | 6 (8.3) | 0.73 (0.08–6.55) | 0.781 | 0.90 (0.08–10.00) | 0.930 |
| Walk | 7 (43.8) | 30 (41.7) | 1.09 (0.37–3.25) | 0.879 | 1.35 (0.43–4.28) | 0.612 |
| Exercise in the classroom (spontaneously) | 3 (18.8) | 26 (36.1) | 0.41 (0.11–1.57) | 0.192 | 0.45 (0.12–1.75) | 0.248 |
| Exercise in the classroom (watching it on television) | 2 (12.5) | 18 (25.0) | 0.43 (0.09–2.07) | 0.292 | 0.50 (0.10–2.50) | 0.397 |
| Frequency of exercise ^a^ |  |  |  |  |  |  |
| None | 0 (0) ^d^ | 0 (0) ^d^ | 1 (reference) ^f^ |  | 1 (reference) ^f^ |  |
| 1–2 days | 0 (0) | 7 (13.2) |  |  |  |  |
| 3–5 days | 2 (18.2) | 21 (39.6) |  |  |  |  |
| Almost every day | 9 (81.8) | 25 (47.2) | 5.04 (0.99–25.60) | 0.051 | 3.79 (0.71–20.30) | 0.120 |
| Duration of exercise ^a^ |  |  |  |  |  |  |
| < 30 minutes | 5 (45.5) | 29 (54.7) | 1 (reference) |  | 1 (reference) |  |
| 30–60 minutes | 2 (18.2) | 16 (30.2) | 0.73 (0.13–4.17) | 0.719 | 0.50 (0.07–3.53) | 0.484 |
| > 60 minutes | 4 (36.4) | 8 (15.1) | 2.90 (0.63–13.40) | 0.173 | 1.92 (0.29–12.90) | 0.500 |
| Working on cognitive training ^b^ |  |  |  |  |  |  |
| No | 5 (41.7) | 24 (41.4) | 1 (reference) |  | 1 (reference) |  |
| Yes | 7 (58.3) | 34 (58.6) | 0.99 (0.28–3.49) | 0.985 | 0.91 (0.23–3.55) | 0.888 |
| Frequency of cognitive training ^c^ |  |  |  |  |  |  |
| None | 0 (0) ^e^ | 0 (0) ^e^ | 1 (reference) ^f^ |  | 1 (reference) ^f^ |  |
| 1–2 days | 4 (50.0) | 21 (60.0) |  |  |  |  |
| 3–5 days | 2 (25.0) | 7 (20.0) |  |  |  |  |
| Almost every day | 2 (25.0) | 7 (20.0) | 1.38 (0.23–8.36) | 0.725 | 1.43 (0.21–9.55) | 0.713 |

Data presented as mean ± standard deviation or number (%). Propensity score was used as an adjustment covariate.

^a^ There were 5 non-responders in the decline group, and 19 non-responders in the non-decline group

^b^ There were 4 non-responders in the decline group, and 14 non-responders in the non-decline group

^c^ There were 8 non-responders in the decline group, and 37 non-responders in the non-decline group

^d^ It is presumed that this item was ignored because participants have already answered on it in the question about “Type of exercise”

^e^ It is presumed that this item was ignored because participants have already answered on it in the question about “Working on cognitive training”

^f^ Reference was the total value of “none,” “1–2 days,” and “3–5 days”

OR, Odds ratio; CI, Confidence interval
